# Identification of GGC Repeat Expansions in *ZFHX3* Among Chilean Movement Disorder Patients

**DOI:** 10.1101/2025.03.17.25323863

**Authors:** Paula Saffie-Awad, Abraham Moller, Kensuke Daida, Pilar Alvarez Jerez, Zhongbo Chen, Zachary B. Anderson, Mariam Isayan, Kimberly Paquette, Sophia B Gibson, Madison Fulcher, Abigail Miano-Burkhardt, Laksh Malik, Breeana Baker, Paige Jarreau, Henry Houlden, Mina Ryten, Bida Gu, Mark JP Chaisson, Danny E. Miller, Pedro Chaná-Cuevas, Cornelis Blauwendraat, Andrew B. Singleton, Kimberley J. Billingsley

**Affiliations:** Clínica Santa María, Santiago, Chile; Center for Alzheimer’s and Related Dementias, National Institute on Aging and National Institute of Neurological Disorders and Stroke, National Institutes of Health, Bethesda, MD, USA; Laboratory of Neurogenetics, National Institute on Aging, National Institutes of Health, Bethesda, MD, USA; Department of Neurodegenerative Disease, UCL Queen Square Institute of Neurology, University College London, London, UK; Department of Genetics and Genomic Medicine, Great Ormond Street Institute of Child Health, University College London, London WC1N 1EH, UK 3; NIHR Great Ormond Street Hospital Biomedical Research Centre, University College London, London WC1N 1EH, UK; Division of Genetic Medicine, Department of Pediatrics, University of Washington, Seattle, Washington, USA; Department of Neurology and Neurosurgery, National Institute of Health, Yerevan, Armenia; Department of Genome Sciences, University of Washington, Seattle, Washington 98195, USA; Department of Neuromuscular Disease, Queen Square Institute of Neurology, UCL, London, UK; Department of Genetics and Genomic Medicine, Great Ormond Street Institute of Child Health, UCL, London, UK; NIHR Great Ormond Street Hospital Biomedical Research Centre, UCL, London, UK; UK Dementia Research Institute at the University of Cambridge, Cambridge, UK; Department of Clinical Neurosciences, School of Clinical Medicine, University of Cambridge, Cambridge, UK; Department of Quantitative and Computational Biology, University of Southern California, Los Angeles, CA, USA; Department of Laboratory Medicine and Pathology, University of Washington, Seattle, Washington 98195, USA; Brotman Baty Institute for Precision Medicine, University of Washington, Seattle, Washington 98195, USA; Centro de Trastornos del Movimiento, Facultad de Ciencias M édicas, Universidad de Santiago de Chile, Santiago, Chile

## Abstract

**Background:** Hereditary ataxias are genetically diverse, yet up to 75% remain undiagnosed due to technological and financial barriers. A pathogenic *ZFHX3* GGC repeat expansion was recently linked to spinocerebellar ataxia type 4 (SCA4), characterized by progressive ataxia and sensory neuropathy, with all reported cases in individuals of Northern European ancestry.

**Methods:** We performed Oxford Nanopore Technologies (ONT) genome long-read sequencing (>115 GB per sample) on a total of 15 individuals from Chile; 14 patients with suspected hereditary movement disorders and one unrelated family member. Variants were identified using PEPPER-Margin-DeepVariant 0.8 (SNVs), Sniffles 2.4 (SVs), and Vamos 2.1.3 (STRs). Ancestry was inferred using GenoTools with reference data from the 1000 Genomes Project, Human Genome Diversity Project, and an Ashkenazi Jewish panel. Haplotype analysis was conducted by phasing SNVs within *ZFHX3*, and methylation profiling was performed with modbamtools.

**Results:** We identified *ZFHX3* GGC repeat expansions (47–55 repeats) in four individuals with progressive ataxia, polyneuropathy, and vermis atrophy. One case presented parkinsonism–ataxia, expanding the phenotype. Longer expansions correlated with earlier onset and greater severity. Hypermethylation was detected on the expanded allele, and haplotype analysis linked ultra-rare *ZFHX3* variants to distant Swedish ancestry.

**Conclusion:** This is the first report of SCA4 outside Northern Europe, confirming a shared founder haplotype and expansion instability. The presence of parkinsonism broadens the clinical spectrum. Comprehensive genetic testing across diverse populations is crucial, and long-read sequencing enhances diagnostic yield by detecting repeat expansions and SNVs in a single assay.

## Introduction

Hereditary ataxias, like many other rare neurological disorders, are challenging to diagnose due to their extensive genetic and clinical heterogeneity. These conditions can result from a wide range of genetic variant types, including short tandem repeats (STRs), single nucleotide variants (SNVs), and structural variants (SVs) such as large insertions, deletions, and other complex rearrangements, making comprehensive molecular diagnosis complex. Despite the availability of targeted gene panels, diagnostic yields remain around 75% ^1^ and can be even lower in resource-limited settings. For example, a specialized center in Chile reported a diagnostic rate of only 23% in patients presenting with ataxia^2^. These gaps underscore the need for continued refinements in both clinical and molecular evaluation strategies.

Among the more than 50 recognized spinocerebellar ataxias (SCAs), SCA4 was first mapped to chromosome 16q22.1 over 25 years ago in a large Utah-based family of Swedish ancestry ^3^. Its underlying genetic etiology was recently identified, as an uninterrupted GGC repeat expansion in the final exon of *ZFHX3* gene, possibly linked to a single founder Northern European haplotype ^4,5^. Notably, this expansion has not been identified in the non-European population ^6,7^ despite screening in Japanese ^8^ and Brazilian cohorts ^9^. Expanded alleles typically range from 42 to 74 repeats, while normal alleles are generally shorter (14–31 repeats) and often contain interruptions by other sequences. Alleles in the 32–41 range have uncertain clinical significance ^10^. Clinically, SCA4 is characterized by a combination of cerebellar ataxia and sensory axonal neuropathy, frequently accompanied by dysautonomia and oculomotor abnormalities ^11^. A notable anticipation phenomenon correlated with repeat size, with a typical age of onset between 30 and 50 years^4–6^. Unlike more frequent ataxias—such as SCA2 and SCA3, SCA4’s full phenotypic spectrum, prevalence in non-European populations, and molecular mechanisms are still being defined.

In this study, we performed Oxford Nanopore Technologies (ONT) genome long-read sequencing on 15 individuals, including two multiplex families and eleven index cases, with suspected hereditary movement disorders—ataxia, ataxia-parkinsonism, parkinsonism, or atypical parkinsonism. We identified SCA4-related heterozygous *ZFHX3* expansions in three unrelated Chilean families, suggesting that SCA4 may be more geographically and ethnically diverse than previously recognized. Here, we present these findings and emphasize the advantages of long-read sequencing for accurately diagnosing SCA4 and other neurodegenerative diseases.

## Methods

### Patients and Participants

This study was approved by the local ethics committee in Chile, ensuring compliance with ethical research standards. Written informed consent was obtained from all participants. Peripheral venous blood samples were collected and stored at −80 °C to preserve the integrity of genetic material for subsequent analyses. A total of 15 samples were collected, from two multiplex families and eleven index cases, with suspected hereditary movement disorders— ataxia, ataxia-parkinsonism, parkinsonism, or atypical parkinsonism **(Supplementary Table 1)**.

### Data generation

The DNA protocols are publicly available on protocols.io (DOI: 10.17504/protocols.io.n92ldmx3ol5b/v1) ^12^. In brief, DNA was extracted from whole blood using the QIAamp Blood Midi Kit (Qiagen), following the spin-column protocol. Whole blood samples, collected in EDTA tubes, were equilibrated to room temperature prior to processing. DNA extraction was performed according to the manufacturer’s protocol, with minor modifications to optimize yield and purity. The extracted DNA was quantified using the Qubit Fluorometer (Thermo Fisher Scientific) with the Qubit dsDNA Broad Range Assay Kit, ensuring high-quality DNA suitable for downstream applications. On average, the protocol yielded 20–30 µg of DNA per sample The samples were then size selected with Sage Science’s Blue Pippin following the 0.75 Agarose Dye-Free 10 kb High Pass Plus Marker U1 protocol. An average of 3.56 µg of DNA was loaded into size selection for each sample. After size selection, the average peak size was 27.8kb. Libraries were constructed using an SQK-LSK114 ONT kit (https://www.protocols.io/view/processing-frozen-archival-human-dna-samples-for-l-5jyl82morl2w/v1) using the same modifications as above however where 1.3-2.5 µg of DNA was used as starting input. PromethION sequencing (MinKNOW version 24.02.10) was performed as per manufacturer’s guidelines with minor adjustments, 20 fmol of the library was loaded onto each primed R10.4.1 flow cell. Each sample required at least two or three loads to achieve > 115GB total data output over 72 hours.

### Data processing

All samples were basecalled and aligned on the NIH HPC Biowulf cluster. Basecalling was performed using dorado v0.7.1 on HPC nodes with 2 NVIDIA V100X GPUs (super-accurate basecalling model - dna_r10.4.1_e8.2_400bps_sup@v5.0.0, 5mCG/5hCG methylation calling) and samples were mapped to the GRCh38 human genome using Minimap2 v2.28 with the map-ont preset. We called SNVs, SVs, and STRs from alignments against GRCh38 with a set of variant detection tools. SNVs were called with Clair3 v1.0.10V and identified with PEPPER-Margin-DeepVariant 0.8 ^13^). SVs were called and merged amongst individuals with Sniffles 2.4^14^ and then split into harmonized per individual VCFs using bcftools 1.21 plugin +split. STRs were called with Vamos 2.1.3^15^. For ancestry analysis, SNVs were assessed using reference data from the 1000 Genomes Project, the Human Genome Diversity Project, and an Ashkenazi Jewish panel, analyzed with GenoTools^16,17^.

In order to compare haplotypes, we took the SNV calls for our samples and looked for the six ultra-rare SNVs coming from the Swedish ancestry founder effect as reported in Chen et al^18^. We then plotted the haplotypes for visual comparison using the following script: https://github.com/zanderson82/SNP-Haplotype-Plotting/tree/main.

### Methylation analysis

To look at methylation patterns, we used modbamtools v0.4.8 (https://www.biorxiv.org/content/10.1101/2022.07.07.499188v1) to generate plots for our expansion carriers and a control. We provided the haplotagged bams from PEPPER-Margin-DeepVariant as input, and the Gencode v38GRCh38 gtf for the gene tracks. Additionally, we added the –hap option to split out the methylation frequency by haplotype.

### Data sharing and code availability

The data supporting the findings of this study are available upon reasonable request due to ethical and legal restrictions related to patient confidentiality. Summary statistics and relevant scripts used for data analysis are publicly available in the https://github.com/molleraj/CARDlongread-chile-data-processing associated with this study. Due to privacy concerns, individual-level genetic and clinical data cannot be shared publicly but can be accessed through data-sharing agreements with the corresponding author upon reasonable request and approval from the relevant ethics committees.

## Results

### Data generation and analysis

We faced significant challenges implementing high-molecular-weight DNA protocols in Chile. These protocols typically require advanced expertise, costly equipment, and complex logistics, including dry ice shipping. To address these obstacles, we developed and publicly shared a cost-effective wet-lab DNA extraction protocol tailored for resource-limited settings, available on the protocols.io platform (https://www.protocols.io/view/protocol-purification-of-dna-from-whole-blood-usin-c7ypzpvn)^12^. DNA was extracted from frozen blood in Chile, with subsequent processing and sequencing performed at the NIH. This workflow generated high-quality ONT whole-genome long-read sequencing data (average N50: 21 kb; 130 GB per flow cell) for under $1,000 per sample **(Supplementary Table 2)**. This dataset facilitated comprehensive analyses of SNVs, SVs, and repeat expansions. A detailed overview of the workflow used for these analyses is presented in **Figure 1a**. Ancestry analysis predicted all the Chilean samples to be Latino/admixed American by Genotools (**Supplementary Figure 1**).

**Figure 1.**
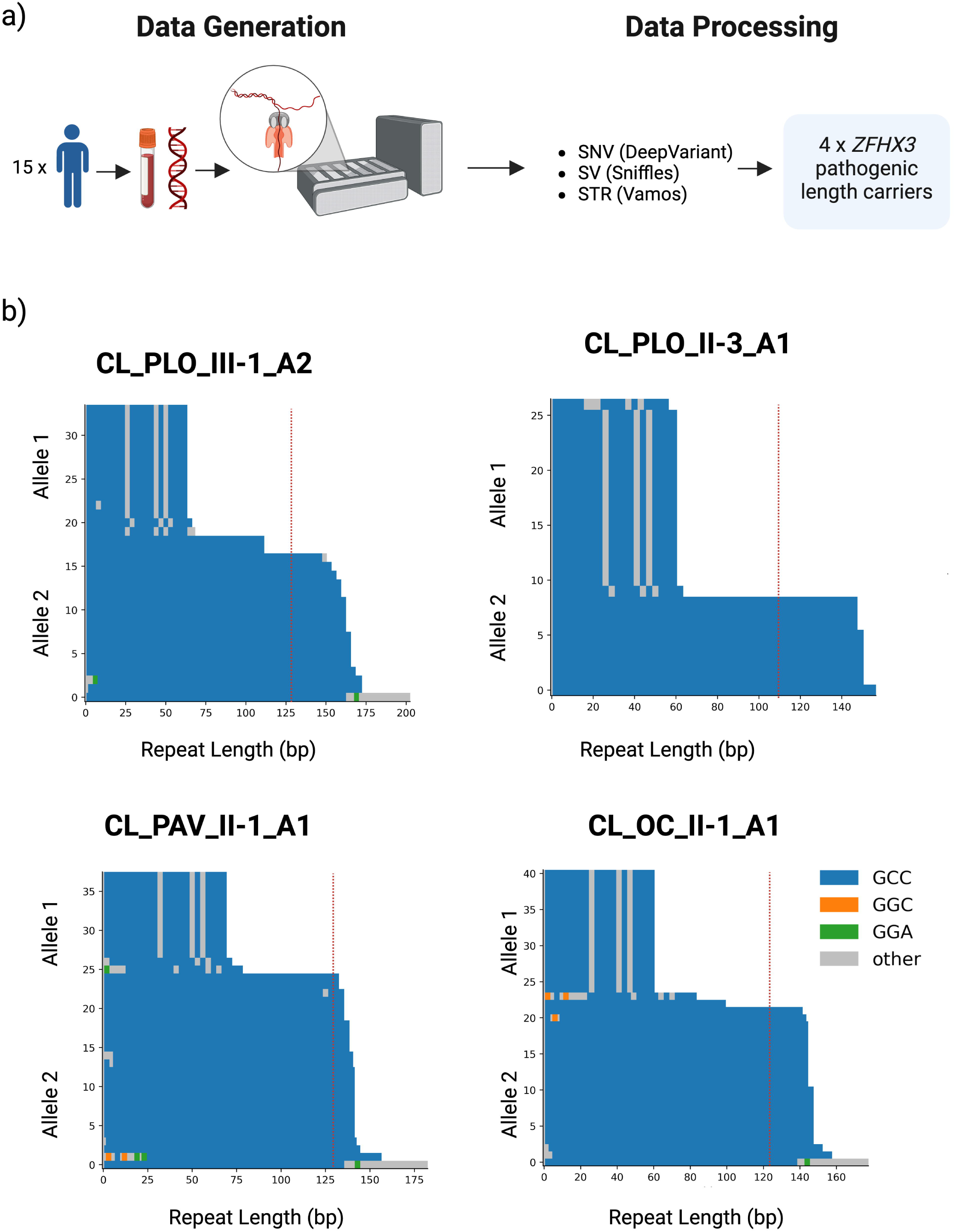
**a)** Schematic overview of the study design. **b)** Waterfall plots displaying ONT long-read sequencing data for the four predicted *ZFHX3* GGC repeat carriers. The red dotted line marks the pathogenic threshold. Created using BioRender.com.

### Identifying four carriers of the GGC *ZFHX3* expansion

Repeat expansions, such as those in *HTT* (CAG repeats in Huntington’s disease), *ATXN1* and *ATXN2* (CAG repeats in spinocerebellar ataxias), and *FXN* (GAA repeats in Friedreich’s ataxia), are well-established causes of neurodegenerative disorders ^19,20^. Using the *vamos* tool, we assessed the lengths of a catalog of known pathogenic STRs. Among the cohort, we identified four patients with pathogenic-length expansions of the recently described *ZFHX3* GGC repeat, associated with SCA4 (**Figure 1.b)**. No other pathogenic-length STRs were detected in the remaining patients. **Supplementary Table 3** provides a summary of the STR lengths determined from long-read sequencing for all tested pathogenic loci. Additionally, no known pathogenic SNVs or SVs were identified in the four *ZFHX3* carriers. An inverse relationship was observed between repeat length and age at onset, with longer expansions associated with earlier disease manifestation. While this trend was not statistically significant (R² = 0.54, p = 0.26) **(Supplementary Figure 2**), it aligns with previous reports ^10^.

To date, the SCA4 expansion has only been identified in individuals who carry ultra-rare SNVs linked to a distant common founder event in Sweden. Here, we analyzed phased SNVs surrounding the repeat expansion and identified four of the six ultra-rare SNVs previously reported in individuals with SCA4 by Figueroa et al.^4^ and Chen et al^18^

.Notably, these SNVs were all absent in non-carriers **(Figure 2, Supplementary Table 4)**.

**Figure 2:**
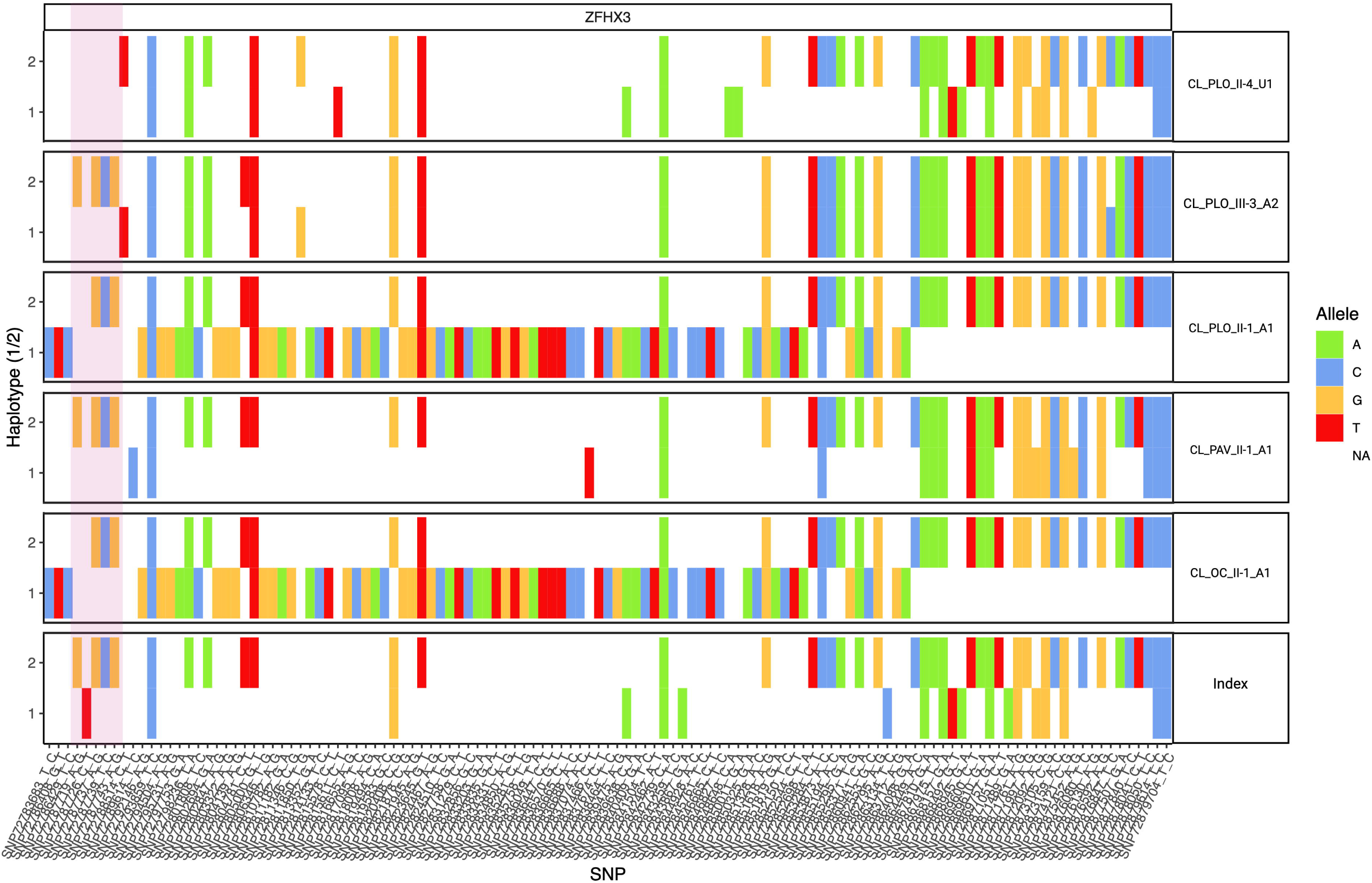
Haplotype analysis of *ZFHX3* GGC repeat expansion carriers. Out of the six rare SNVs reported as part of the distance common founder event, four were found in our samples within the repeat region (Highlighted by the pink box) and compared to the Utah index patient from Chen et al^18^. These SNVs are missing in the unaffected Chilean individual.

### Methylation analysis

Chen et al. previously reported hypermethylation associated with the SCA4 expansion, prompting us to investigate methylation patterns in our cohort^18^. Consistent with their findings, haplotype-specific methylation calling around the *ZFHX3* GGC repeat expansion showed hypermethylation around the STRs compared with the non-expanded allele overlapping with the last exon of *ZFHX3*. In contrast, samples without the expansion showed hypomethylation in this region **(Figure 3)**. This differential methylation pattern present at the expanded allele suggests that the presence of the GGC repeat could have downstream effects on *ZFHX3* expression through epigenetic regulation.

**Figure 3.**
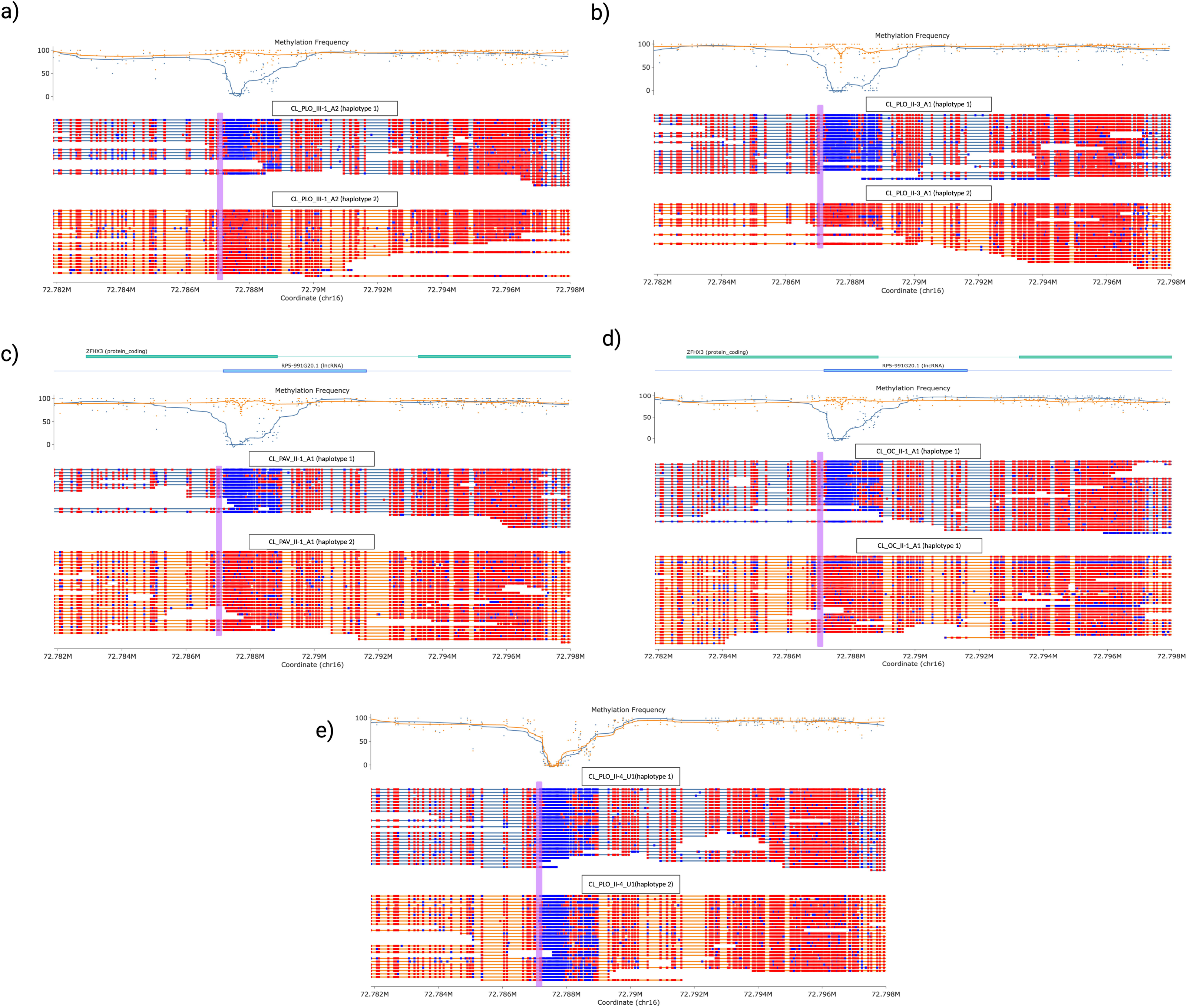
Haplotype specific Differential methylation around the expansion. **a-d)** Modbamtools plots of methylation frequency for the four heterozygous expansion carriers. Methylation frequency is plotted at the top where haplotype 1 (blue) represents the non-expanded allele and haplotype 2 (orange) represents the expanded allele. The *ZFHX3* gene track is overlaid at the top, showing the last two exons. Haplotype-specific reads are shown at the bottom with blue sections of the reads denoting hypomethylation and red sections of the reads denoting hypermethylation. Purple box indicates repeat region. For all four carriers, the expanded allele is hypermethylated compared to the non-expanded allele. **e)** Modbamtools plot of an unaffected related individual, homozygous non- repeat carrier, showing that hypomethylation in this region is expected under normal conditions.

### Clinical characteristics and phenotype of *ZFHX3* carriers

All four *ZFHX3* carriers initially presented with generalized ataxia without vertigo or cerebellar atrophy, accompanied by various neurological and non-neurological features, including chronic cough. Notably, one patient (CL_OC_II- 1_A1) progressed to a rapidly evolving parkinsonism–ataxia syndrome. A detailed summary of each patient’s clinical presentation is provided in **Table 1**.

CL_PLO_III-1_A2, a female with a 10-year disease duration, carries 55 CAG repeats. She exhibited a classic ataxic phenotype (SARA 10) without dysarthria, motor neuron involvement, or cognitive impairment. Electromyography (EMG) confirmed polyneuropathy, and her only non-neurological symptom was chronic cough. Magnetic Resonance Imaging (MRI) showed mild cerebellar atrophy. CL_PLO_II-3_A1, a male with a 9-year disease duration, carries 49 CAG repeats. His presentation included severe ataxia (SARA 28), dysarthria, and upper motor neuron signs, such as spasticity and a positive Babinski sign. He demonstrated mild cognitive decline (MoCA 21) and polyneuropathy with hypopalesthesia. Additional features included bladder incontinence and REM sleep behavior disorder (RBD). Neuroimaging revealed cerebellar vermis atrophy. CL_PAV_II-1_A1, a female with an 8-year disease duration, carries 47 CAG repeats. She presented with gait ataxia (SARA 19), dysarthria and sensory polyneuropathy. No vertigo, upper motor neuron involvement, or cognitive decline was observed. Her only additional complaint was chronic cough. MRI demonstrated upper vermis and upper spinal cord atrophy. There was no reported family history of neurological disorders. CL_OC_II-1_A1, a male with a 5-year disease duration, carries 49 CAG repeats. He presented with a rapidly progressive parkinsonism–ataxia phenotype characterized by severe ataxia, spasticity, a positive Babinski sign, and moderate-to-severe polyneuropathy confirmed by EMG. Significant cognitive decline was observed alongside other features, including gaze palsy, severe bladder incontinence, constipation, and myoclonus. MRI revealed cerebellar vermis atrophy.

This analysis highlights the overlap of clinical features associated with *ZFHX3* expansions with more common ataxias, such as *RFC1*-related disorders and Friedreich’s ataxia making diagnosis challenging when relying solely on the clinical phenotype and testing for a single expansion.

### *ZFHX3* Repeat Length Distribution Across Diverse Cohorts

Although no large-scale long-read sequencing datasets are currently available from individuals of Chilean ancestry, ancestry analysis of our samples aligned with Admixed American (AMR) populations. Chileans exhibit a complex genetic background primarily composed of Native American and European ancestry, with the Native American component closely related to Andean indigenous groups such as the Mapuche. The European ancestry in Chileans predominantly derives from Southern Europe, with some Northern European influence, while a smaller but detectable contribution of African ancestry reflects historical migration patterns in Latin America ^21^.

To contextualize our findings, we leveraged data from two large long-read sequencing resources: the CARD Long-Read Initiative and the 1000 Genomes (1000G) Long-Read Project. From the CARD Long-Read Initiative, we analyzed 205 control samples of European ancestry from the North American Brain Expression Consortium (NABEC) cohort and 133 control samples of African and African-admixed ancestry from the Human Brain Collection Core (HBCC) cohort^22^. Additionally, we screened 100 samples of mixed ancestry from the 1000G Long-Read Project^23^. To ensure methodological consistency with the Chilean samples, all repeat lengths were analyzed using *vamos*. In the CARD dataset, *ZFHX3* GGC repeat lengths ranged from 11 to 24 in the NABEC cohort and from 18 to 23 in the HBCC cohort. Similarly, in the 1000G dataset, repeat lengths ranged from 18 to 29 **(Figure 4).** To date, this represents the most comprehensive analysis of *ZFHX3* repeat length variability using long-read sequencing. These results establish a robust reference for *ZFHX3* repeat length variation across ancestrally diverse control cohorts, providing a critical framework for comparison with patient samples. Further, our findings support the repeat size threshold of ≥42 proposed by Wallenius et al., reinforcing its potential role in SCA4 pathogenicity^5^.

**Figure 4.**
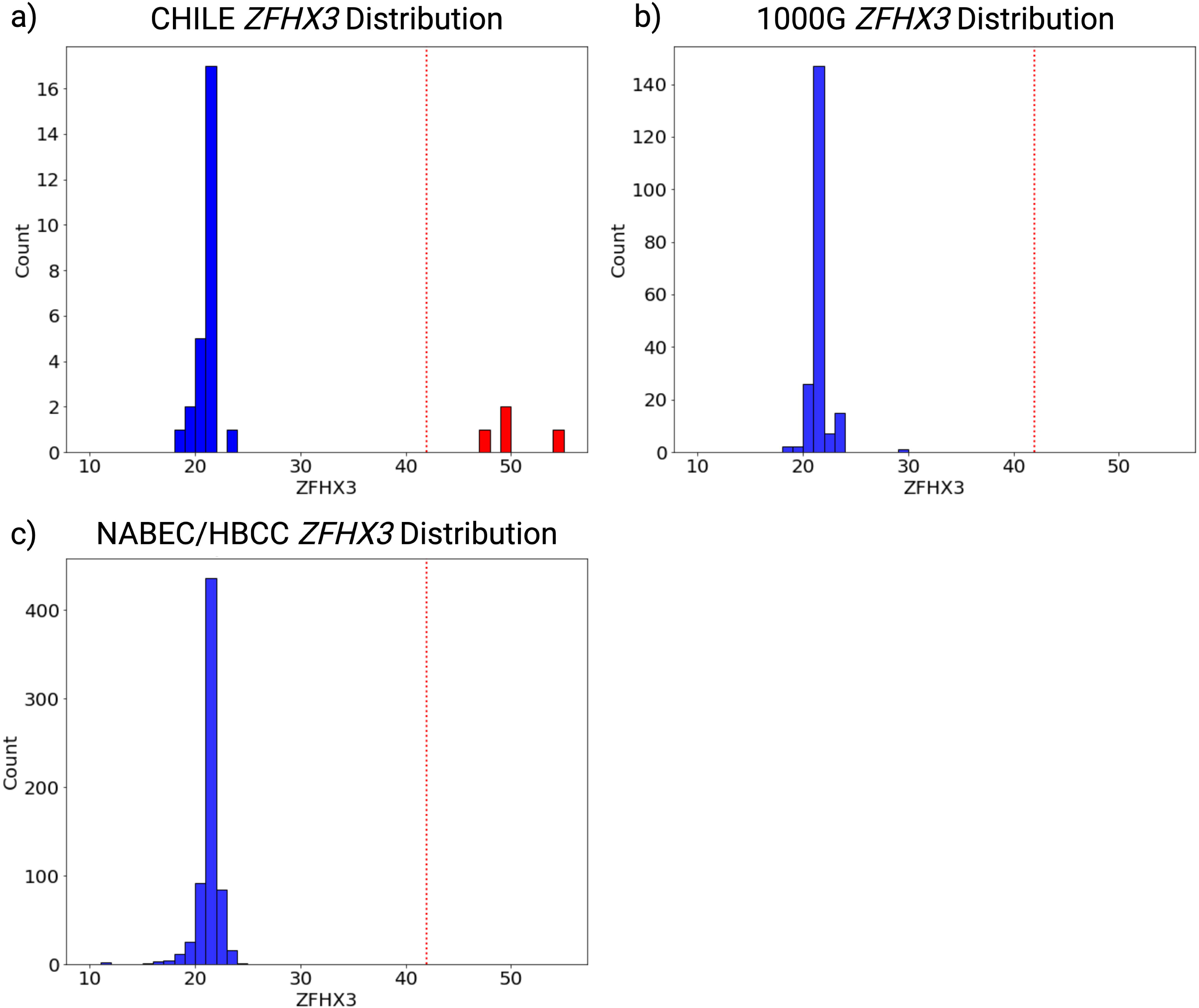
**a)** Distribution of ZFHX3 GGC repeat lengths in the Chilean samples (n=15), read indicates pathogenic length carrier. **b)** Distribution of ZFHX3 GGC repeat lengths in the 1000G control cohort (n=100) comprising individuals of mixed ancestry. **c)** Distribution of ZFHX3 GGC repeat lengths in the NABEC/HBCC control cohort (n=338) comprising individuals of European and African and African-admixed ancestry. The red dotted line marks the pathogenic threshold.

## Discussion

Our study represents the largest analysis of the SCA4 STR using long-read sequencing across diverse ancestries, incorporating NABEC (n=205), HBCC (n=133), and a subset from the 1000 Genomes Project (n=100). Despite limited South American datasets, this work expands the known geographic distribution of SCA4, reporting the first confirmed cases outside Northern Europe and reinforcing the need for broader population studies^24^

A key finding is the intergenerational expansion of the *ZFHX3* repeat, with an increase from 49 to 55 units in an affected family. This supports the anticipation phenomenon, a well-documented feature in repeat expansion disorders. Notably, a recent study demonstrated a strong inverse correlation between repeat length and age at onset, with longer repeats associated with earlier disease presentation (CL_PLO_III-1_A2) and greater severity (CL_OC_II-1_A1). Our findings align with this, suggesting that repeat instability may influence disease progression in SCA4^10^.

Clinically, all affected individuals exhibited ataxia, polyneuropathy, upper motor neuron signs, and dysautonomia, consistent with SCA4 ^25^. However, we identified parkinsonism in one patient, a feature not previously linked to this disorder. Given its occurrence in other SCAs (SCA2, SCA3, SCA6, SCA8, SCA17)^26,27^, and the the overlapping neurodegenerative processes affecting multiple brain regions ^28^ this suggests that SCA4 may have a broader clinical spectrum rather than this being a coincidental finding. Further studies, including neuroimaging and functional assays, are needed to clarify this association.

At the molecular level, we confirmed hypermethylation of the expanded *ZFHX3* allele, suggesting an epigenetic role in disease pathogenesis. Similar methylation changes in *C9orf72*-related ALS/FTD ^29,30^ and Fragile X syndrome^31^ have been associated with transcriptional dysregulation, highlighting the need for further investigation into the functional consequences of methylation in SCA4. Additional follow-up studies are required to determine how the expansion affects *ZFHX3* expression and function. Consistent with previous reports, we identified a common Swedish founder haplotype, estimated to have originated approximately 2,200 years ago. The detection of this haplotype outside Northern Europe, including in our Chilean cohort, suggests a wider historical dispersal of the pathogenic expansion^6^.

Despite these findings, limitations remain. The small sample size underscores the need for larger, multi-ethnic studies, and the lack of genomic data from diverse populations limits full assessment of *ZFHX3* variability. Expanding sequencing efforts in underrepresented populations is critical for refining our understanding of SCA4. While long-read sequencing is needed to capture the full spectrum of structural variants, repeat expansions, and complex regions, it can be resource-intensive. An efficient approach would be to implement adaptive sampling^6^, enabling targeted enrichment of regions associated with ataxias to comprehensively detect all variant types including SNVs, STRs, and SVs, while optimizing sequencing efficiency and reducing costs. This targeted sequencing strategy could effectively address the current limitations of ataxia panels, providing a comprehensive solution for genetic diagnosis.

In summary, this study expands the knowledge of SCA4 by confirming a shared founder haplotype, documenting repeat expansion instability, and identifying a potential link to parkinsonism. Addressing current limitations through broader genetic studies and improved diagnostic access will be key to advancing understanding and clinical management of SCA4.

## Acknowledgements

We sincerely thank the patients and their families for their participation in this study. Their invaluable contributions make this research possible.

We also acknowledge the support of Oxford Nanopore Technologies staff in generating this dataset, in particular A. Markham, J. Anderson, and C. Vacher.

This work was supported in part by the Intramural Research Program of the National Institute on Aging (NIA) and the Center for Alzheimer’s and Related Dementias (CARD), within the Intramural Research Program of the NIA and the National Institute of Neurological Disorders and Stroke (ZIANS003154, ZIAAG000538), National Institutes of Health (AG000538). Computational resources were provided by the NIH HPC Biowulf cluster (https://hpc.nih.gov).

Additional support was provided by the National Institutes of Health (DP5OD033357, R01HG011649, 5T32HG000035-29), as well as grants from the Michael J. Fox Foundation, the Medical Research Council, the Wellcome Trust, and the National Institute for Health and Care Research (UCL/UCLH Biomedical Research Centre).

## Authors’ Roles

P.S.A., A.M., K.D., P.A.J., C.B., Z.C.: Design, execution, analysis, writing, editing of final version of the manuscript.

K.P., L.M., B.B., M.F., M.I., A.M.B., A.B.S., Z.B.A., D.E.M: Execution, analysis, editing of final version of the manuscript.

H.H., M.R., B.G., M.J.P., P.J. S.B.G: : Analysis, editing of final version of the manuscript.

K.J.B.: Supervision, design, execution, analysis, writing, editing of final version of the manuscript.

## Financial Disclosure of all authors (for the preceding 12 months)

This work was supported in part by the Intramural Research Program of the National Institute on Aging (NIA) and the Center for Alzheimer’s and Related Dementias (CARD), within the Intramural Research Program of the NIA and the National Institute of Neurological Disorders and Stroke (ZIANS003154, ZIAAG000538, AG000538), National Institutes of Health (NIH). Computational resources were provided by the NIH HPC Biowulf cluster (https://hpc.nih.gov).

<u>Note</u>: Individuals affiliated with CARD or the Laboratory of Neurogenetics at NIH are marked with an asterisk (*) below, as this is their funding source. None of the authors have received consultancies, honoraria, hold intellectual property rights, or receive royalties

- *Paula Saffie-Awad* – Employment: Clínica Santa María, Santiago, Chile. Grant: Michael J. Fox Foundation. Advisory Boards: Biogen.
- *Pilar Alvarez Jerez* * – Employment: Laboratory of Neurogenetics, National Institute on Aging, NIH, Bethesda, MD, USA; Department of Neurodegenerative Disease, UCL Queen Square Institute of Neurology, University College London, UK.
- *Cornelis Blauwendraat, Andrew B. Singleton*\* – Employment: Center for Alzheimer’s and Related Dementias, National Institute on Aging and National Institute of Neurological Disorders and Stroke, NIH, Bethesda, MD, USA; Laboratory of Neurogenetics, National Institute on Aging, NIH, Bethesda, MD, USA.
- *Abraham Moller* \**, Kimberly Paquette* *, Laksh Malik*, Breeana Baker*, Paige Jarreau*, Kimberley J. Billingsley* – Employment: Center for Alzheimer’s and Related Dementias, NIH, USA.
- *Kensuke Daida* \**, Madison Fulcher* *, Abigail Miano-Burkhardt* – Employment: Laboratory of Neurogenetics, NIH, Bethesda, MD, USA.
- *Zhongbo Chen* – Employment: UCL Queen Square Institute of Neurology, UK; Great Ormond Street Institute of Child Health, UCL, UK.
- *Zachary B. Anderson* – Employment: University of Washington, Seattle, WA, USA.
- *Sophia B. Gibson* – Employment: Division of Genetic Medicine, Department of Pediatrics, University of Washington, Seattle, WA, USA; Department of Genome Sciences, University of Washington. **Grant Support:** NIH grant 5T32HG000035-29.
- *Danny E. Miller* – Employment: Division of Genetic Medicine, Department of Pediatrics, University of Washington, Seattle, WA, USA; Department of Laboratory Medicine and Pathology and the Brotman Baty Institute for Precision Medicine, University of Washington. **Stock Ownership:** MyOme. **Advisory Boards:** Oxford Nanopore Technologies (ONT), Basis Genetics. **Partnerships:** ONT, Pacific Biosciences, Basis Genetics. **Grant Support:** NIH DP5OD033357.
- *Mariam Isayan* – Employment: National Institute of Health, Yerevan, Armenia.
- *Henry Houlden* – Employment: UCL Queen Square Institute of Neurology, UK. **Grants:** Michael J. Fox Foundation, MRC, Wellcome Trust, NIHR UCL/UCLH BRC.
- *Mina Ryten* – Employment: UCL, UK; University of Cambridge, UK.
- *Bida Gu, Mark JP* – Employment: University of Southern California, USA. **Grant Support:** NIH R01HG011649.

## Tables and figures Legends

**Table 1. Clinical characteristics of the four *ZFHX3* GGC repeat carriers.** Abbreviations: SARA, Scale for the Assessment and Rating of Ataxia; MRI, magnetic resonance imaging; RBD, Rapid Eye Movement sleep behavior disorder, PKN, for parkinsonism, EMG, Electromyography.

**Supplementary Figure 1.** Scatter plot of principal components 1 and 2 from the Ancestry analysis Cluster plot of PC1 and PC2 of the ancestry analysis showed the Chilean samples clustering close to Latino/admixed American (AMR). AAC; African American/Afro-Caribbean, AFR; African, AJ; Ashkenazi Jewish, AMR; Admixed American, CAS; Central Asian, EAS; Eastern Asian, EUR; European, FIN; Finnish, MDE; Middle Eastern, SAS; South Asian.

**Supplementary figure 2.** Inverse Correlation Between *ZFHX3* GGC Repeat Length and Age at Onset in SCA4 Patients. A negative trend is observed, but it is not statistically significant (R² = 0.54, p = 0.26).

## Supplementary Tables

**Supplementary Table 1. Clinical phenotypes of the individuals studied.** Sample ID based on their country (CL for Chile), family ID, generation within the pedigree (Roman numerals: I, II, III, etc.), individual order within that generation (numbered from left to right in the pedigree), and affected status (A for affected, U for unaffected).

**Supplementary Table 2. Overview of long-read sequencing data for the studied.** N50 represents the read length at which 50% of total bases are in reads of that length or longer. Yield (Gb) indicates total bases sequenced. Mean coverage refers to the average sequencing depth.

**Supplementary Table 3. Overview of lengths per allele of known pathogenic expansion loci.** The table includes chromosome (#chr) position, affected gene, associated disease, pathogenic repeat number, and repeat lengths for each individual.

**Supplementary Table 4. Summary results of haplotype analysis of six ultra-rare SNVs associated with the Swedish ancestry founder effect** ^4,18^. Genotypes for the *ZFHX3* carriers at six SNV on chromosome 16 (hg38). Shared haplotypes are highlighted. *Abbreviations:* SNV, single nucleotide variant; hg38, human genome assembly GRCh38.

## Notes

The authors declare no financial or non-financial conflicts of interest related to this manuscript.

### Competing Interest Statement

The authors declare that this work was supported in part by the Intramural Research Program of the National Institute on Aging (NIA) and the Center for Alzheimers and Related Dementias (CARD) within the Intramural Research Program of the NIA and the National Institute of Neurological Disorders and Stroke (ZIANS003154, ZIAAG000538, AG000538), National Institutes of Health (NIH), with computational resources provided by the NIH HPC Biowulf cluster. Authors affiliated with CARD or the Laboratory of Neurogenetics at NIH received funding from these institutions.
D.E.M. holds stock in MyOme, serves on the advisory boards of Oxford Nanopore Technologies (ONT) and Basis Genetics, and has partnerships with ONT, Pacific Biosciences, and Basis Genetics. P.S.A. serves on an advisory board for Biogen. Several authors received grant support, including P.S.-A. (Michael J. Fox Foundation), S.B.G. (NIH 5T32HG000035-29), D.E.M. (NIH DP5OD033357), H.H. (Michael J. Fox Foundation, MRC, Wellcome Trust, NIHR UCL/UCLH BRC), and B.G. and M.J.P. (NIH R01HG011649).

### Clinical Protocols

https://www.protocols.io/view/protocol-purification-of-dna-from-whole-blood-usin-c7ypzpvn

https://github.com/molleraj/CARDlongread-chile-data-processing

### Funding Statement

This study was supported in part by the Intramural Research Program of the National Institute on Aging (NIA) and the Center for Alzheimers and Related Dementias (CARD), within the Intramural Research Program of the NIA and the National Institute of Neurological Disorders and Stroke (ZIANS003154, ZIAAG000538), National Institutes of Health (AG000538).
Additional support was provided by the National Institutes of Health (DP5OD033357, R01HG011649, 5T32HG000035-29), as well as grants from the Michael J. Fox Foundation, the Medical Research Council, the Wellcome Trust, and the National Institute for Health and Care Research (UCL/UCLH Biomedical Research Centre)

### Author Declarations

The Ethics Committee of Centro de Trastornos del Movimiento (CETRAM) gave ethical approval for this work. Written informed consent was obtained from all participants

