## Supplementary figures and images for "Identification of GGC Repeat Expansions in *ZFHX3* Among Chilean Movement Disorder Patients"

### Supplementary Figure 1

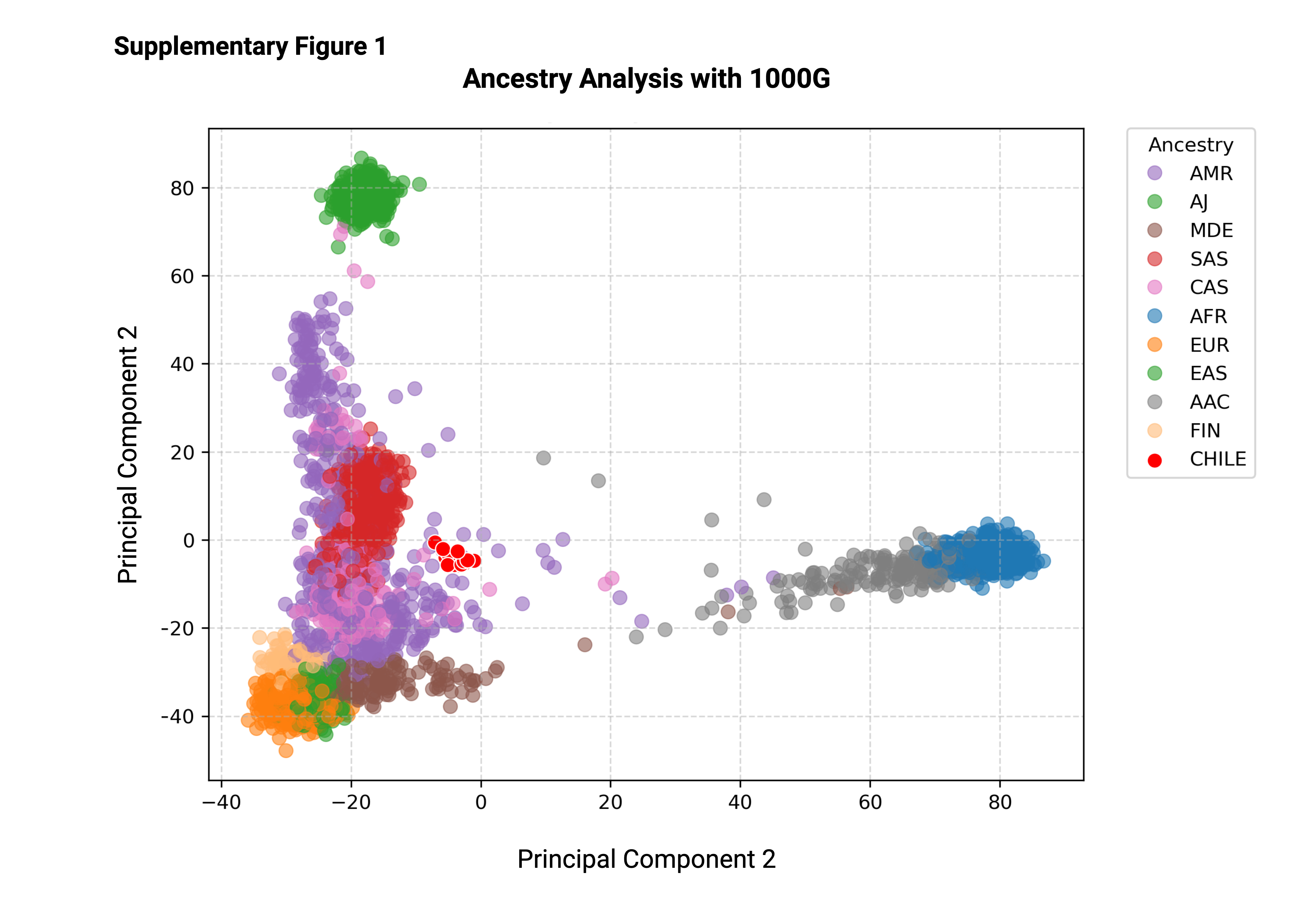

### Supplementary Figure 2

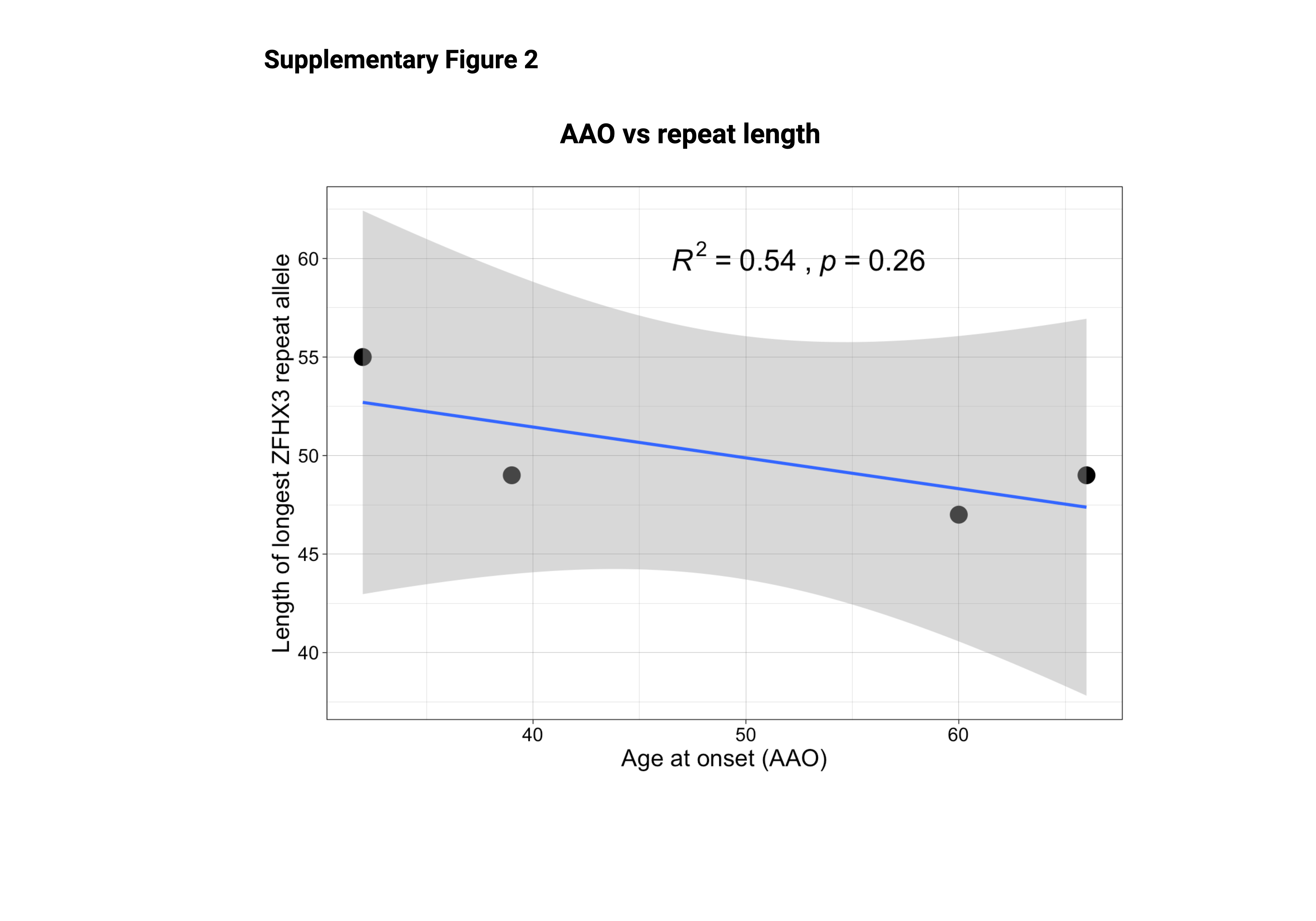
